# Sex disparities in in-hospital outcomes after percutaneous coronary intervention (PCI) in patients with acute myocardial infarction and a history of coronary artery bypass grafting (CABG): A nationwide inpatient sample-matched analysis (2016–2019)

**DOI:** 10.1101/2024.03.11.24304146

**Authors:** Rui Yan, Xueping Ma, Bo Shi, Congyan Ye, Shizhe Fu, Kairu Wang, Haohong Qi, Mingzhi Cui, Ru Yan, Shaobin Jia, Guangzhi Cong

## Abstract

**Background:** The role of sex disparities in in-hospital outcomes after percutaneous coronary intervention (PCI) for acute myocardial infarction (AMI) in patients with a history of coronary artery bypass grafting (CABG) remains underexplored. This study aimed to identify sex disparities in in-hospital outcomes after PCI in patients with AMI and a history of CABG.

**Methods:** Using the National Inpatient Sample database, we identified patients hospitalized for AMI with a history of CABG who underwent PCI between 2016 and 2019. The primary outcome was in--hospital mortality, and the secondary outcomes were the length of hospital stay and hospitalization costs. 1:1 propensity score matching was used to minimize standardized mean differences of baseline variables and compare in--hospital outcomes.

**Results:** In total, 75,185 weighted hospitalizations of patients who underwent PCI were identified, with 25.2% being female patients. Compared with male patients, female patients exhibited elevated risks of in-hospital mortality (3.72% vs. 2.85%, p = 0.0095), longer length of stay (4.64 days vs. 3.96 days, p < 0.001), and higher hospitalization costs ($112,594.43 vs. $107,020.54, p = 0.0019). After propensity score matching, female patients had higher in-hospital mortality rates than male patients (3.81% vs. 2.89%, p = 0.028). Multivariable logistic regression (adjusted odds ratio [aOR]: 1.48; 95% confidence interval [CI]: 1.14–1.92) and propensity score matching (aOR: 1.34; 95% CI: 1.03–1.73) showed a consistently higher risk of in-hospital mortality among female patients than among male patients. Female patients aged >60 years were more vulnerable to in-hospital mortality than were their male counterparts (3.06% vs. 4.15%, p = 0.0003, aOR: 1.55; 95% CI: 1.18–2.04).

**Conclusions:** Female patients who underwent PCI for AMI with a history of CABG had a higher in-hospital mortality rate, which was particularly evident among older patients aged >60 years. Therefore, sex- and age-specific investigations and interventions are required to reduce disparities within this high-risk population.

## Introduction

Coronary artery bypass grafting (CABG) is the most commonly performed cardiac surgical procedure in adults [1] . However, despite advances in pharmacological and surgical techniques, saphenous vein graft (SVG) failure rates remain significant [2, 3]. Furthermore, the development of atherosclerosis, thrombosis, and calcification in native coronary arteries is accentuated by bypass grafting [4, 5]. Consequently, patients with a history of CABG remain at risk for recurrent ischemic episodes in the months and years after surgery. This risk is primarily attributed to SVG occlusion and the progression of native coronary artery disease [6, 7]. Moreover, these patients frequently require additional revascularization because they experience recurring angina symptoms or present with acute coronary syndrome [8, 9]. The technique and strategy of further revascularization for this unique population remain challenging issues in therapeutic interventions for coronary heart disease (CAD). In these patients, simple pharmacological therapy adjustments frequently fail to control myocardial ischemia. Due to anatomical alterations, tissue adhesions, and the source of the bridging arteries, secondary thoracotomies for repeat CABG are not often recommended. Therefore, in patients with a susceptible anatomy, percutaneous coronary intervention (PCI) is the preferred revascularization strategy [10–12].

Female patients were more likely to undergo urgent CABG, but they still have a higher risk of post-procedural complications [13]. Previous studies have shown that the incidence of recurrent MI in female patients is higher than that in male patients after CABG surgery. Consequently, female patients often require further revascularization, such as PCI [14–16]. The female sex is seen as a risk factor for negative outcomes after PCI [17]. Although this disparity has repeatedly drawn attention, it has primarily been attributed to variations in risk factor profiles between men and women, because it is diminished or eliminated in a multivariable analysis that controls for baseline confounders [18, 19]. Currently, no existing report in the literature has compared sex discrepancies in in-hospital outcomes after PCI among patients with acute myocardial infarction (AMI) who have a history of CABG. Consequently, debate persists owing to a lack of evidence. Thus, further research is required to elucidate sex differences in in-hospital outcomes following PCI in this patient population.

This study aimed to identify sex disparities in in-hospital outcomes after PCI in patients with AMI who have a history of CABG, using a nationally representative cohort. The hypothesis posited that the in-hospital mortality rate among female patients would be higher than that among male patients.

## Materials and methods

### Data source

This study utilized the National (Nationwide) Inpatient Sample (NIS), which is the largest all-payer database of hospital inpatient stays in the United States. The NIS is a component of the Healthcare Quality and Utilization Project (HCUP), sponsored by the Agency for Healthcare Research and Quality. It provides discharge information from a 20% stratified sample of community hospitals [20]. The NIS includes approximately 7 million unweighted records and 35 million weighted hospital encounters annually. National estimates can be measured using weighted data. The information provided for each discharge includes patient demographics, primary payer status, socioeconomic status, hospital features, primary diagnosis, up to 24 secondary diagnoses, and procedural diagnoses. The HCUP-NIS does not record information about specific individual patients; rather, it records all data pertaining to a particular admission. Because the data within this database is de-identified and accessible to the public, the requirement for informed consent was waived. This study was approved by the Institutional Review Board of Ningxia Medical University and complied with the Strengthening the Reporting of Observational Studies in Epidemiology (STROBE) reporting guidelines [21].

### Study design and population

Between January 1, 2016, and December 31, 2019, all hospitalizations of adults aged ≥18 years with a primary discharge diagnosis of AMI and a history of CABG, along with a procedural diagnosis of PCI, were included. These hospitalizations were stratified by sex. The International Classification of Diseases-10 Clinical Modification (ICD-10) and Clinical Classification Software codes were used to identify the clinical outcomes, procedures, and patient comorbidities (S1 Table). The 31 AHRQ Elixhauser comorbidity assessments currently in use were used to identify additional comorbidities [22] (S2 Table). To consider any variations at the hospital level, hospital-related variables, such as hospital bed size, region, and location/teaching status, were studied. The exclusion criteria were as follows: (1) receipt of CABG during the index admission (n = 97), (2) history of prior PCI between 2016 and 2019 (n = 7,759), (3) age <18 years (n = 1), and (4) missing demographic data (n = 1089) (Fig 1).

**Fig 1.**
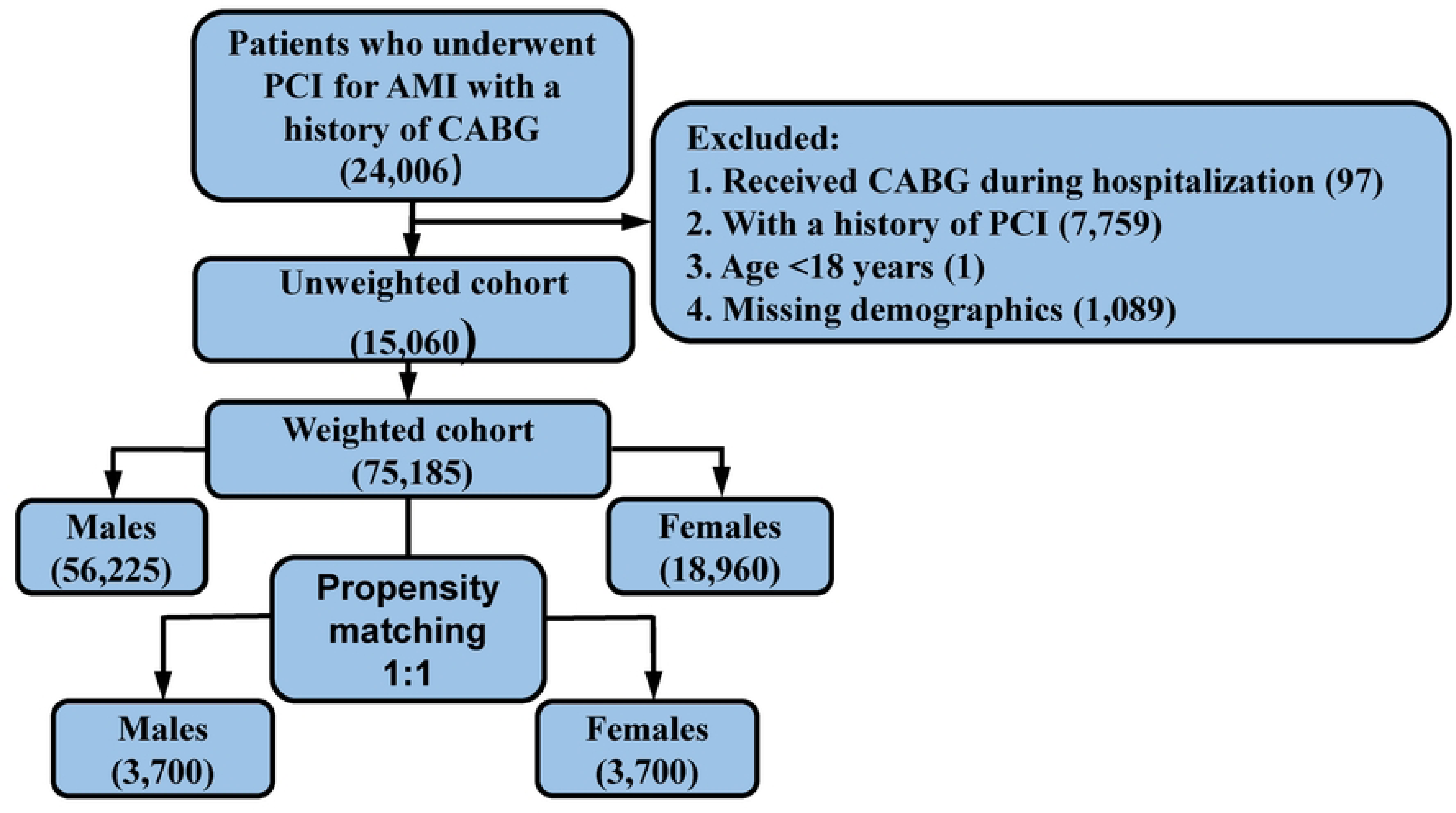
Flowchart of patient selection. CABG, coronary artery bypass grafting; PCI, percutaneous coronary intervention; AMI, acute myocardial infarction

### Outcomes

The primary outcome of interest was in-hospital mortality. The secondary outcomes were the length of hospital stay, hospitalization costs, and difference in in-hospital mortality between male and female hospitalized patients according to age.

### Statistical analysis

All variables are presented as weighted national estimates in accordance with HCUP-NIS guidance [23]. This was performed in accordance with the survey analysis strategy, using HOSP_NIS as a clustering variable and NIS_STRATUM to account for the different strata in the NIS design, as suggested by the Agency for Healthcare Research and Quality Methods series. For continuous variables, data are presented as survey-weighted mean (95% confidence intervals [CI]), and the p-value was calculated using survey-weighted linear regression. For categorical variables, data are reported as survey-weighted percentage (95% CI), and the *P*-value was calculated using the survey-weighted chi-square test.

Logistic regression models were used to assess the relationship between sex and in-hospital mortality. The adjusted OR (aOR) and 95% CI were computed after applying the multivariate-adjusted models. These confounders were selected based on their connections with in-hospital mortality, a change-in-effect estimate exceeding 10%, or a regression coefficient p-value of <0.1 with respect to the relationship between sex and in-hospital mortality [24]. Given the nonrandomized nature of the study design, 1:1 propensity score matching was used to match male and female patients to assess the impact of sex on outcomes and mitigate confounding and selection biases via multivariable logistic regression. Matching variables are presented in S3 Table, with a matching tolerance set at 0.05. Standardized mean differences for all baseline variables were calculated to assess the balance of baseline characteristics before and after matching. A standardized mean difference <0.10 was regarded as a good balance of the baseline variable. The relationship between age and in-hospital mortality among male and female patients was explored using smooth curve fitting and subgroup analysis.

The R statistical software package (http://www.R-project.org; The R Foundation) and EmpowerStats (http://www.empowerstats.com; X&Y Solutions, Inc., Boston, MA, USA) were utilized for all analyses. Statistical significance was set at p < 0.05 (two-sided).

## Results

### Patient characteristics

We identified 15,060 unweighted patients who underwent PCI for AMI with prior CABG in the NIS database between January 1, 2016, and December 31, 2019. This patient population comprised 11,261 (74.77%) and 3,799 (25.23%) male and female patients, respectively. After applying discharge weight, this represented 56,225 and 18,960 male and female patients, respectively. Compared with male patients, female patients were more likely to be Black, have a lower socioeconomic status, and have Medicare/Medicaid as their primary payer (all p < 0.001). A higher prevalence of obesity, diabetes, hypothyroidism, chronic pulmonary disease, valvular disease, depression, and fluid and electrolyte disorders (all p < 0.001) was observed among female patients than among male patients. Smoking, alcohol abuse, prior AMI, dyslipidemia, congestive heart failure, hypertension, ventricular fibrillation, and intracardiac thrombus were more common among male patients (all p < 0.05). Overall, female patients had a higher burden of Elixhauser comorbidities (comorbidity index >4, 33.10% vs. 27.11%, p < 0.001) (Table 1). PSM well balanced the baseline variables, with almost all the standardized mean differences <0.1 (S4 Table).

**Table 1.**
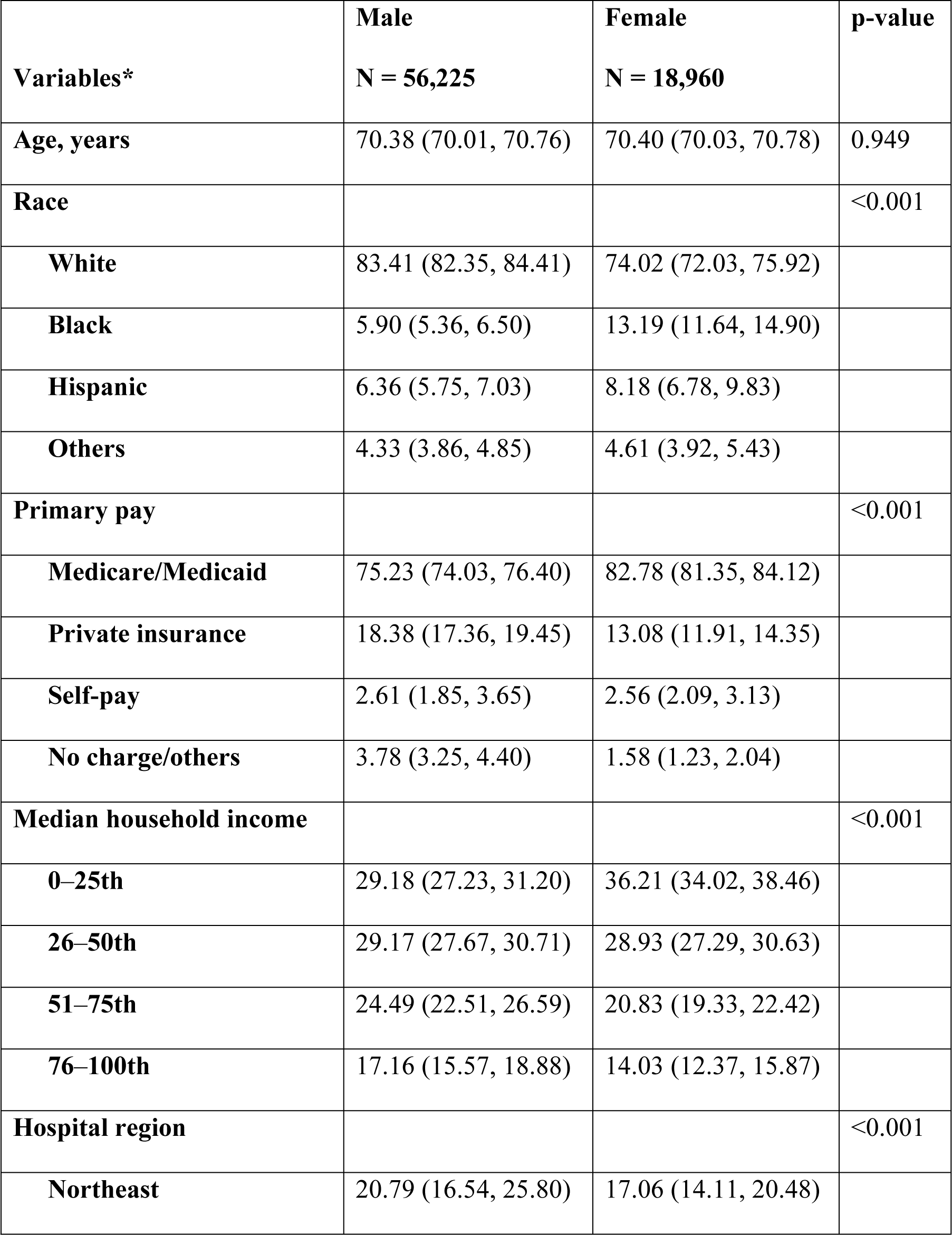
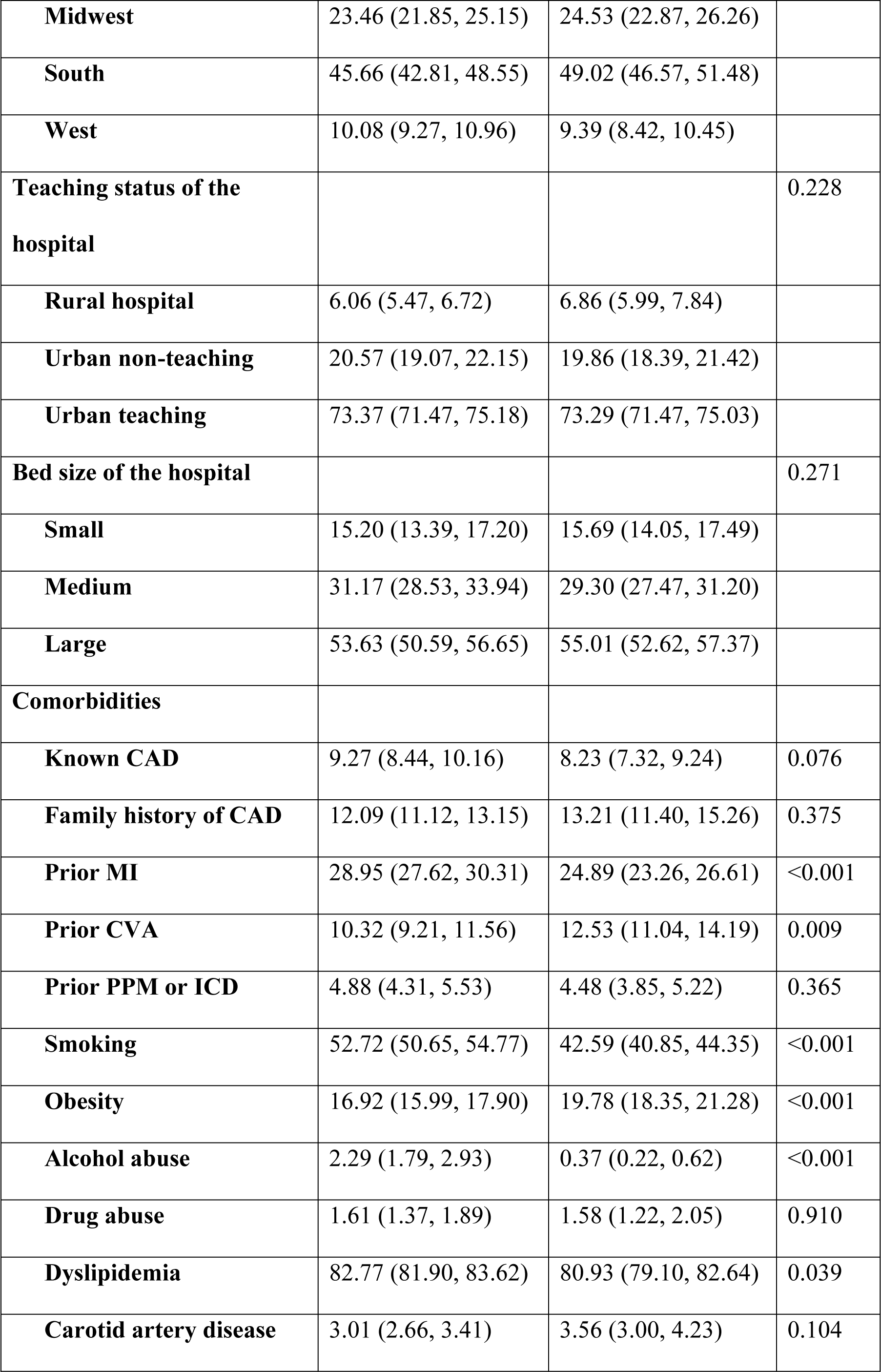
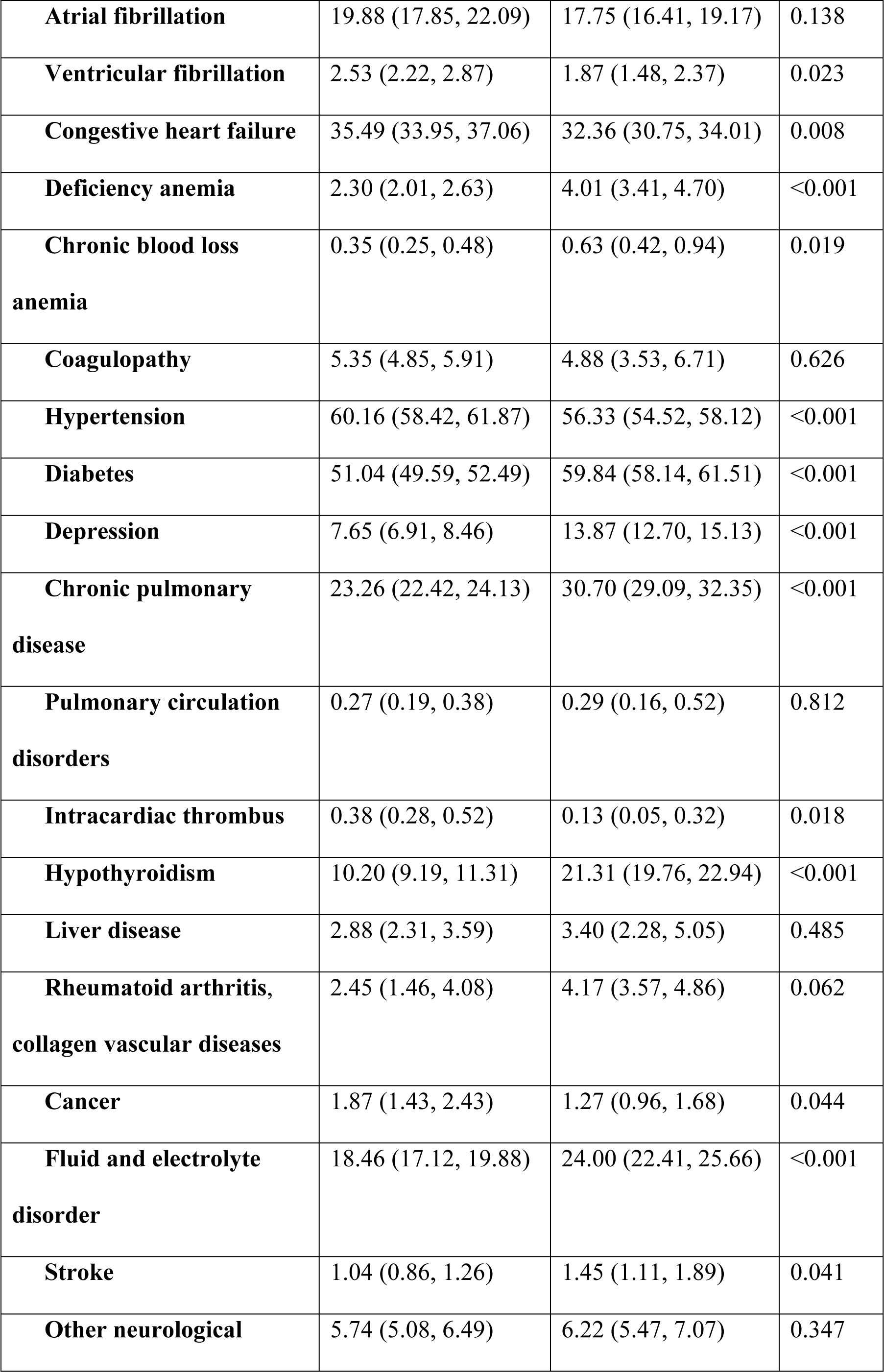
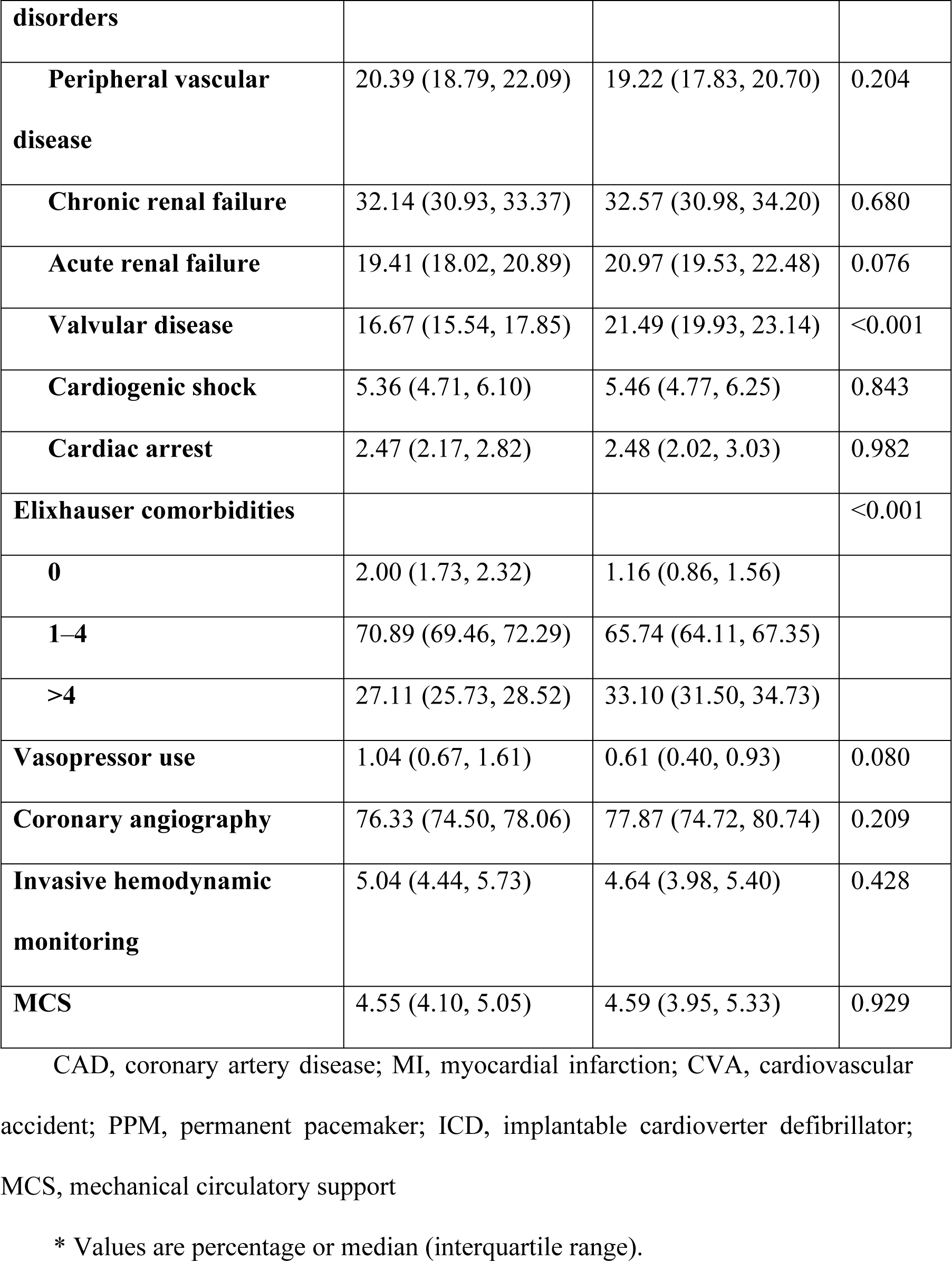
Baseline characteristics of female versus male patients in the cohort before propensity score matching (weighted)

### Outcomes

During the hospital stay, in the overall cohort, female patients had a higher prevalence of in-hospital mortality (3.72% vs. 2.85%, p = 0.009), longer length of stay (4.64 days vs. 3.96 days, p < 0.001), and higher hospitalization costs ($112,594.43 vs $107,020.54, p = 0.002) than did male patients. After propensity score matching, female patients had a higher in-hospital mortality rate and longer length of stay than did male patients (3.81% vs. 2.89%, p = 0.028 and 4.61 ± 4.76 days vs. 4.02 ± 4.35 days, p < 0.001, respectively), while the relationship between sex and hospitalization costs was no longer significant ($113,797.03 ± 96,086.46 vs. $111,768.45 ± 92,179.28, p = 0.354) (Tables 2 and 3). Multivariable logistic regression (aOR: 1.48; 95% CI: 1.14–1.92) and propensity score matching (aOR: 1.34; 95% CI: 1.03–1.73) showed a consistently higher risk of in-hospital mortality among female patients than among male patients (Table 4).

**Table 2.** Comparison of in-hospital outcomes between female and male patients in the overall cohort.

|  | Male, N = 56225 | Female, N = 18960 | p-value |
| --- | --- | --- | --- |
| **In-hospital mortality (%)** | 2.85 (2.52, 3.23) | 3.72 (3.14, 4.39) | 0.009 |
| **Length of stay, days** | 3.96 (3.85, 4.06) | 4.64 (4.44, 4.84) | <0.001 |
| **Total hospitalization costs, \$** | 107020.54<br>(103859.25, 110181.83) | 112594.43<br>(108917.49, 116271.37) | 0.002 |

Values are percentage or median (interquartile range).

**Table 3.** Comparison of in-hospital outcomes between female and male patients in the matched cohort.

|  | Male, N = 3700 | Female, N = 3700 | p-value |
| --- | --- | --- | --- |
| **In-hospital mortality (%)** | 2.89 | 3.81 | 0.028 |
| **Length of stay, days** | 4.02 ± 4.35 | 4.61 ± 4.76 | <0.001 |
| **Total hospitalization costs, \$** | 111,768.45±92,179.28 | 113,797.03 ± 96,086.46 | 0.354 |

Values are percentage or mean ± SD

**Table 4.** In-hospital mortality of multivariable logistic regression and propensity-matched multivariate logistic regression.

|  | aOR (95% CI) | p-value |
| --- | --- | --- |
| **Overall cohort** | 1.48 (1.14, 1.92) | 0.004 |
| **Matched cohort** | 1.34 (1.03, 1.73) | 0.029 |

Note: aOR (95% CI) P-value

The model was adjusted for the following factors: age; race, primary pay, ZIP income, hospital region, teaching status of the hospital, bed-size of the hospital, smoking, dyslipidemia, family history of coronary artery disease, prior myocardial infarction, atrial fibrillation, congestive heart failure, deficiency anemia, chronic blood loss anemia, chronic pulmonary disease, coagulopathy, depression, diabetes mellitus; hypertension, hypothyroidism, liver disease, fluid and electrolyte disorder, other neurological disorders, pulmonary circulation disorders, chronic renal failure, valvular disease, obesity, cardiogenic shock, stroke, ventricular fibrillation, cardiac arrest, acute renal failure, coronary angiography, invasive hemodynamic monitoring, vasopressor use, mechanical circulatory support.

aOR, adjusted odds ratio; CI, confidence interval

### Comparison of in-hospital mortality according to age

Smooth curve fitting revealed an upward trend in in-hospital mortality in all age groups, with a larger increase occurring among patients aged >60 years. Subgroup analysis revealed that female patients aged >60 years were more vulnerable to in-hospital mortality than were their male counterparts (3.06% vs. 4.15%; aOR: 1.55; 95% CI: 1.18–2.04) (Table 5 and Fig 2).

**Fig 2.**
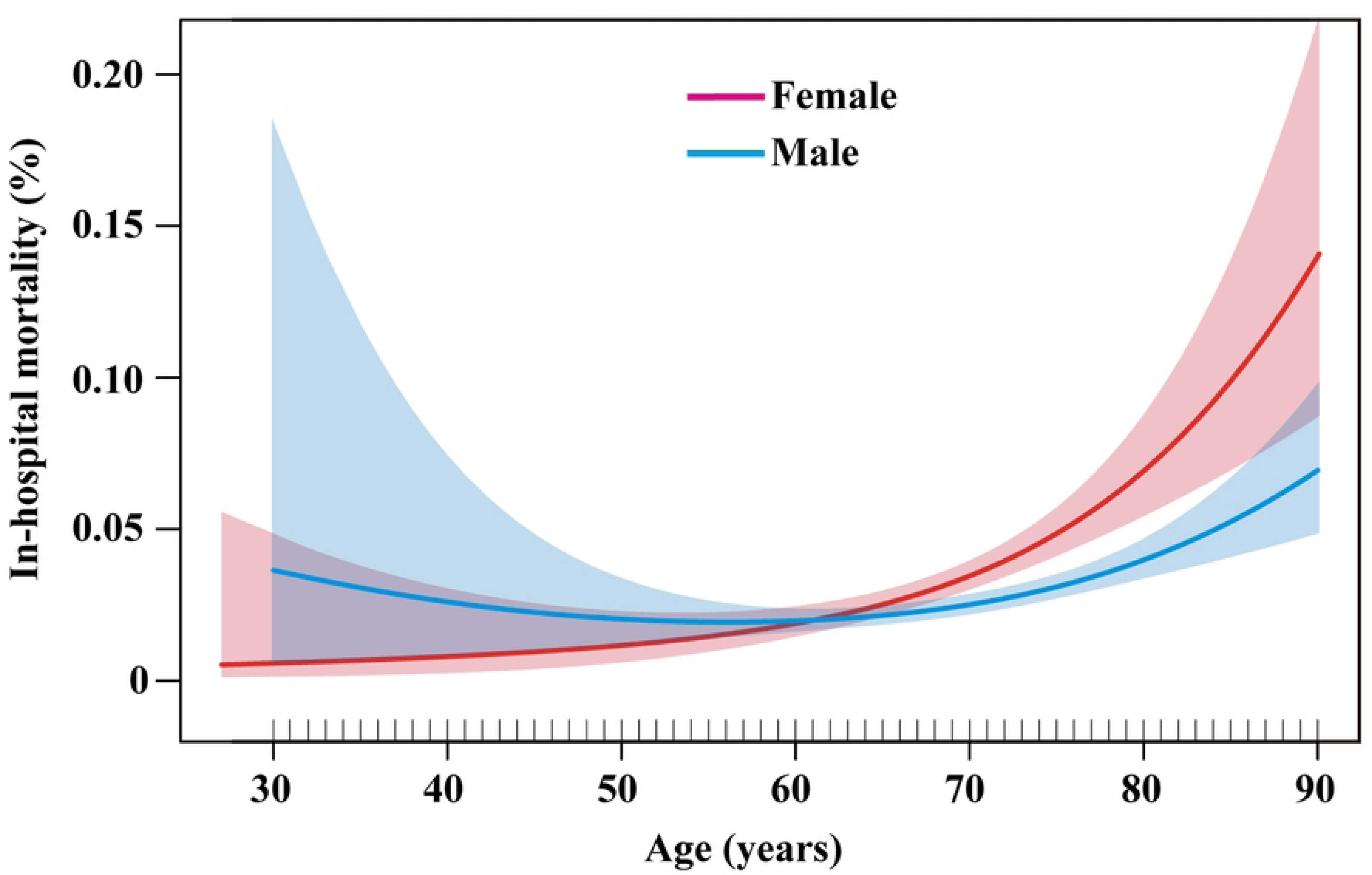
Comparison of in-hospital mortality according to age.

**Table 5.** Subgroup analysis of in-hospital mortality according to age.

|  | Male | Female | aOR (95% CI) | p-value |
| --- | --- | --- | --- | --- |
| **Age ≤ 60 years** | 40/2007<br>(1.99% ) | 12/692<br>(1.73%) | 0.91 (0.36, 2.33) | 0.848 |
| **Age > 60 years** | 283/9254<br>(3.06%) | 141/3107<br>(4.15%) | 1.55 (1.18, 2.04) | 0.002 |

aOR: adjusted odds ratio

The model was adjusted for the following factors: race; primary pay; ZIP income; hospital region; teaching status of the hospital; bed-size of the hospital; smoking; dyslipidemia; family history of coronary artery disease; prior myocardial infarction; atrial fibrillation; congestive heart failure; deficiency anemia; chronic blood loss anemia; chronic pulmonary disease; coagulopathy; depression; diabetes mellitus; hypertension; hypothyroidism; liver disease; fluid and electrolyte disorder; other neurological disorders; pulmonary circulation disorders; chronic renal failure; valvular disease; obesity; cardiogenic shock; stroke; ventricular fibrillation; cardiac arrest; acute renal failure; coronary angiography; invasive hemodynamic monitoring; vasopressor use; mechanical circulatory support.

## Discussion

In this study, we investigated sex disparities in the in-hospital outcomes of patients with AMI who underwent PCI and had a history of CABG. The main findings were as follows: (1) in both the overall and matched cohorts, female patients had a higher in-hospital mortality rate than did male patients and (2) female patients aged >60 years were at greater risk of in-hospital mortality than were their male counterparts.

### Sex differences in in-hospital outcomes in patients who underwent PCI for AMI and had a history of CABG

Numerous reports have explored sex disparities in in-hospital outcomes following revascularization in patients with AMI [17, 18, 25]. However, data on sex differences in recurrent hospitalization and repeat revascularization are limited. Previous studies have shown that both SVG failure and the progression of native artery disease contribute to the need for repeat revascularization following CABG [26]. Compared with male patients, female patients were associated with higher rates of recurrent revascularization due to narrower native coronary arteries and bypass conduits, a higher tendency to spasm, and incomplete revascularization [15, 27-31]. Female patients often prefer receiving PCI over undergoing repeat CABG because of their smaller chest cavities and thinner and smaller arteries, which increase their vulnerability to mechanical complications [32, 33]. Nevertheless, no existing study in the literature has explored sex differences in in-hospital outcomes after PCI in patients with a history of CABG. In our overall and matched cohorts, the risk of in-hospital mortality was higher in female patients than in male patients who underwent PCI.

Several possible explanations can be proffered for the increased risk of in-hospital mortality after PCI in female patients with a history of CABG. First, compared with male patients, female patients with AMI typically seek medical attention later following symptom onset; tend to experience longer delays even after reaching the emergency room, including longer triage and door-to-balloon times; and have lower rates of medical therapy that is guided by guidelines, which may be a factor in poorer outcomes [34, 35]. Other factors contributing to longer delays in seeking medical care among female patients, compared with male patients, include lack of knowledge, less aggressive treatment, fear, humiliation, and earlier incorrect diagnoses of chest discomfort by a medical professional [35–37]. Second, the higher in-hospital mortality rates among female patients may be partially attributed to their greater burden of comorbidities than that among male patients [38, 39]. Consistent with these observations, the current study revealed that female patients were more likely to have comorbidities, including diabetes, renal failure, hypothyroidism, chronic pulmonary disease, valvular disease, and fluid and electrolyte disorders. Even after adjusting for demographics, hospital characteristics, and comorbidities, female patients tended to have a higher in-hospital mortality rate than did their male counterparts. In some circumstances, these differences could be linked to pathophysiological variations. Third, anatomical and pathophysiological differences, such as smaller arterial diameters and worse hemodynamic states in female patients, can cause higher risks of bleeding problems and vascular injury [40, 41] . Additionally, left main lesions in female patients are often ostial and do not require bifurcation stenting. However, they are more frequently severely calcific and necessitate the use of rotational atherectomy [42]. Fourth, according to current recommendations, female patients receive less antiplatelet therapy, fewer statin prescriptions, and fewer referrals to cardiac rehabilitation [43].

### Sex differences in in-hospital mortality were more pronounced among older patients

Notably, this study found that female patients aged >60 years were at greater risk of in-hospital mortality than were their male counterparts. Anderson et al. analyzed outcomes in patients aged >65 years who underwent PCI and showed that female patients had a higher in-hospital mortality rate [44]. However, the study only included patients aged >65 years, whereas the current study enrolled patients of all age groups with a history of CABG. Several factors contribute to a higher in-hospital mortality rate among older female patients.

First, multiple studies have demonstrated that estrogen can reduce myocardial apoptosis, protect myocardial cells, and prevent plaque rupture. After menopause, when the preventive effects of estrogen subside, the risk of CAD in women increases significantly. Hence, female patients presenting with CAD tend to be older than their male counterparts, suffer multiple complications with aging, and have a higher in-hospital mortality rate [45, 46]. Notably, older female patients have significantly higher rates of cardiac hypertrophy, which is a risk factor for mortality [47]. Additionally, the incidence of cerebral hemorrhage and embolic strokes is higher among older female patients [48, 49]. Furthermore, age and female sex are two of the most important characteristics linked to cardiac rupture following myocardial infarction [50]. Second, when compared with the transfemoral method, transradial PCI showed a reduced complication rate and a higher technical success rate in older patients [51]. Notably, older female patients tend to prefer femoral artery access [52]. Finally, older female patients with CAD undergoing PCI were frequently weak and malnourished, and these factors are strongly associated with increased all-cause mortality [53, 54]. Further investigation is required to examine the impact of sex and age on in-hospital mortality and repeat revascularization in patients with a history of CABG.

### Limitations

This study has several limitations. First, although the use of ICD-10 codes has been validated for cardiovascular outcomes research, the NIS is an administrative dataset, which may contain coding errors [55]. Additionally, the NIS lacks clinical details, such as medication, biochemistry, and imaging data. Moreover, ICD-10 diagnosis codes could not be used to determine the extent and severity of AMI. They also did not allow us to evaluate the quantity of revascularized vessels, extent of revascularization, quantity and type of stents, or the total length of the stents. In addition, no long-term follow-up data were available in the NIS dataset. Second, as with all retrospective studies, this study may have been subject to selection and observational biases. Third, due to the vague coding for coronary spasm or spontaneous coronary artery dissection, conditions more commonly found in young female patients presenting with AMI, they could not be determined in this cohort [56]. Finally, we could not assess the details of the progression of native coronary arteries and SVG failure after CABG.

### Strengths

Despite these limitations, to the best of our knowledge, no study has examined sex disparities in patients with AMI and a history of CABG who undergo PCI. The large sample size provided ample statistical power to detect sex disparities in clinical outcomes among the patient groups studied. Furthermore, our data represent the best current evidence in the US population, considering that patients with a history of CABG are frequently excluded from or underrepresented in landmark PCI trials. In addition, multivariate analysis and propensity score matching were performed to minimize confounding and selection biases. Finally, the NIS contains information from more than 7 million hospital stays annually, which can be applied to the entire American population.

### Conclusions

The current study showed that female patients who underwent PCI for AMI with a history of CABG had a higher in-hospital mortality rate, which was more pronounced among older patients aged >60 years. Therefore, sex- and age-specific investigations and interventions are required to reduce disparities within this high-risk population.

## Data Availability

The third party data underlying the results presented in this study are available upon request from the Healthcare Cost and Utilization Project (HCUP) - National (Nationwide) Inpatient Sample (NIS) via their website (https://www.hcup-us.ahrq.gov/nisoverview.jsp). All interested researchers can access the data through HCUP directly. The authors of this study are not permitted to share the data or make it publicly available as per the data use agreement with HCUP. The authors did not have any special access privileges to this data.

## Acknowledgements

We acknowledge the NIS (Nationwide Inpatient Sample) for providing the data.

## Supporting information

**S1 Table. International Classification of Diseases, 10th Revision, Clinical Modification/Procedure Coding System (ICD-10 CM/PCS) Codes.**

**S2 Table. 31 AHRQ Elixhauser comorbidity measures.**

**S3 Table. Matching variables in propensity score analysis.**

**S4 Table: Baseline characteristics of female versus male patients in the cohort after propensity score matching.**

